# Rapid detection of SARS-CoV-2 by pulse-controlled amplification (PCA)

**DOI:** 10.1101/2020.07.29.20154104

**Authors:** Zwirglmaier Katrin, Weyh Maria, Krüger Christian, Ehmann Rosina, Müller Katharina, Wölfel Roman, Stoecker Kilian

## Abstract

In the current pandemic of SARS-CoV-2, rapid identification of infected individuals is crucial for management and control of the outbreak. However, transport of samples, sample processing and RT-qPCR analysis in laboratories are time-consuming. Here we present a nucleic acid-based test format – pulse controlled amplification – that allows detection of SARS-CoV-2 directly from up to eight swab samples simultaneously without the need for RNA extraction within 20 min in a point-of-care setting.

## Introduction

The gold standard for diagnosis of acute COVID-19 is currently RT-qPCR. The method is highly sensitive and can detect fewer than 10 copies of the virus genome ^1^. The workflow from sample to result takes about 4-6 hours and includes extraction of the viral RNA before setting up and running the RT-qPCR reaction. Including the time for sample transportation, in-house laboratory logistics, analysis and communication of test results, sample turnaround times easily add up to 24-48 hours and more. Thus, there is an urgent need to cut down the time of SARS-CoV-2 diagnostics.

PCA (pulse-controlled amplification) is a qPCR-related nucleic acid amplification technology that allows detection of a nucleic acid target within a few minutes ^2^. It does not require RNA extraction from the sample material and the test is performed using a lightweight (< 1 kg) device (Pharos Micro, GNA Biosolutions, Martinsried, Germany) that can run off battery power if necessary. PCA is therefore ideally suited for a point of care setting or for use in a drive-through testing station, thereby obviating sample transport. The technology has already been used successfully for DNA targets. In a proof-of-concept study assays for the detection of MRSA and *Yersinia pestis* were developed ^2^. Both assays allowed detection of the target DNA in under 10 minutes. Furthermore, for *Yersinia pestis* a detection was demonstrated for crude sample material without nucleic acid extraction in a simulated bioterror scenario in the field, with the operators wearing full PPE and the Pharos Micro device running off battery power. With this in mind, we adapted PCA for RNA targets and developed a RT-PCA assay to detect SARS-CoV-2 directly from swab samples without prior RNA extraction. The aim was to establish a detection system for SARS-CoV-2 that is a) faster than RT-qPCR and b) does not rely on a laboratory environment and can therefore be used, e.g., at drive-through or mobile testing stations to provide fast test results and allow immediate action on positive test results.

## Material and Methods

### Sample material

SARS-CoV-2 virus culture, strain IMB-1, was grown on Vero E6 cells. Cell culture supernatant containing viral particles was treated either with 4 volumes of AVL buffer (Qiagen, Hilden, Germany) and 1 volume of ethanol or heated to 80 °C for 10 min to inactivate the virus.

Clinical samples consisted of residual diagnostic sample material, i.e. swab samples in universal transport medium.

### Pulse controlled amplification (PCA) - technology

PCA technology is based on thermocycling using local heating for the denaturation step. The amplification takes place on an array of 75 gold-coated tungsten wires with a diameter of 15 µm, which are immersed in the reaction solution and carry one of the primers. Short electric pulses of a few hundred microseconds heat the wires to roughly 95°C and thereby denature double stranded DNA near the wires. During the brief heating period, the temperature field diffuses only a few microns away from the surface of the wires, thus effectively heating only a minute fraction (<1%) of the reaction volume. After the pulse, the heat dissipates nearly instantaneously into the bulk of the solution. This results in very short amplification cycles of 1.5 - 5 seconds, depending on the assay and target sequence. In the Pharos Micro prototype, wires are embedded in a disposable chip that has eight wells, i.e. eight reactions can be run simultaneously. Amplification of the target sequence is detected in real time via hydrolysis probes, similar to qPCR.

### PCA assay for the detection of SARS-CoV-2

Primers and probe sequences used in this study are listed in table 1. Pharos reaction chips were functionalized with the thiolated reverse primer as described by Müller et al.^2^. Sample material (i.e. swab samples in virus transport medium) was treated either with 4 volumes of AVL buffer (Qiagen, Germany) and 4 volumes of ethanol or simply heated to 80 °C for 10 min without adding any lysis buffer. 45 µl of treated sample material and 15 µl of hybridization buffer (GNA Biosolutions) were added to a well in the chip and incubated for 5 min at ambient temperature to allow hybridization of viral RNA to the reverse primer immobilized on the wires. The liquid was then removed and the well rinsed with 50 µl wash buffer (GNA Biosolutions). Optionally, this initial binding step can be repeated to improve sensitivity of the test. 50 µl of the RT-PCA mastermix (containing 7 mM MgCl_2_, 100 nM E-Sarbeco-P1-Rc, 450 nM E-Sarbeco-F, 1x PCA mastermix/ 0.09 x RT mix/ 1x booster mix [GNA Biosolutions]) were added to each well. PCA settings for the Pharos Micro were 5 min reverse transcription at 55 °C, with a lid temperature of 63 °C and base temperature of 55 °C, followed by a 60 sec thermalizing step (to equilibrate the reaction solution to the combined annealing and elongation temperature of 67 °C base and 74 °C lid). For denaturation, 250 µsec pulses were applied to the array of wires suspended in the solution, with the pulses being repeated every 3 sec for 800 cycles. With these settings, the entire run is completed within 40 minutes. The majority of positive samples are detected within 15 min.

**Table 1.**
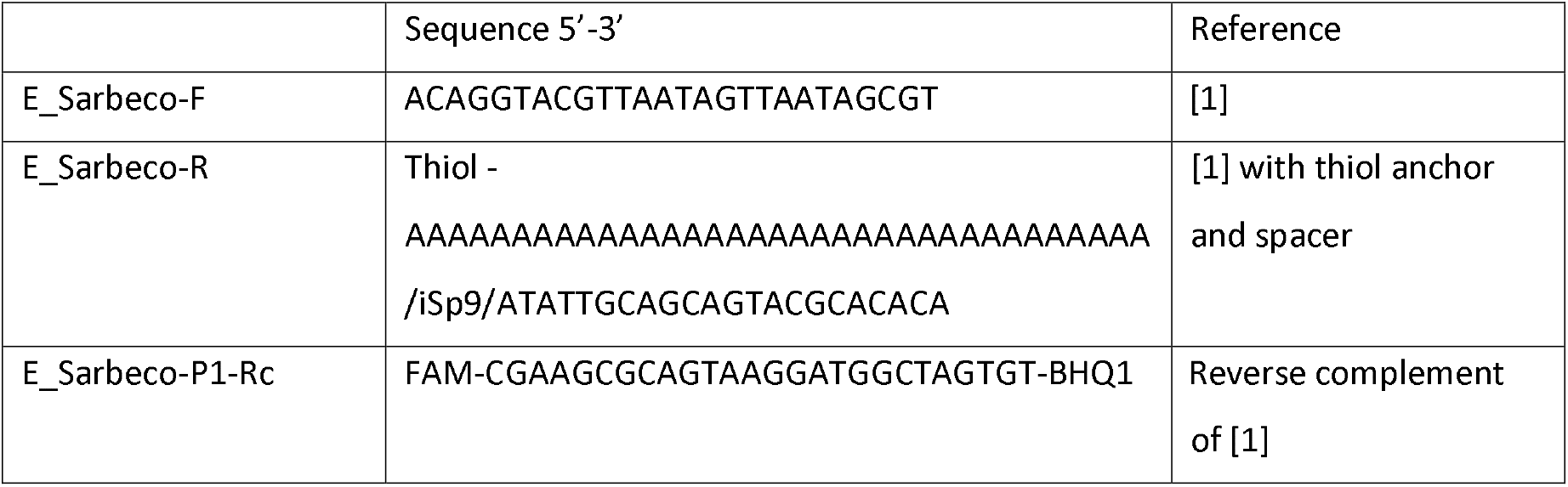
Sequences of primers and probes used in this study. iSp9 is an internal triethylene gycol spacer.

### RT-qPCR

Viral RNA was extracted for RT-qPCR using the QiaAmp Viral RNA extraction kit (Qiagen, Hilden, Germany) following the manufacturer’s protocol.

Primers and probes as well as PCR parameters were used as described by Corman et al. ^1^ The reactions were carried out on a MIC (Magnetic Induction Cycler, BMS, Australia) using Qiagen OneStep RT-PCR reagents.

## Results

We developed a PCA assay for SARS-CoV-2 targeting the E gene. The primer and probe sequences were modified from the assay published by Corman ^1^ to adapt it to the Pharos system, adding a thiolated anchor sequence for the reverse primer to enable it to bind to the gold surface of the wires and using the reverse complement of the published probe sequence (Table 1).

The aim was to arrive at a workflow that allows fast, sensitive and specific detection of SARS-CoV-2 without the need for prior RNA extraction. This is especially important with regard to the global supply shortages in RNA extraction kits seen over the last few months. We used AVL buffer (Quiagen, Hilden Germany) to lyse the viral particles, but did not perform a subsequent complete column-based RNA purification. AVL contains guanidinium thiocyanate, which is a common component of lysis buffers in commercial RNA extraction kits and is known to denature proteins and lyse cells ^3^. However, it is also known to inhibit conventional PCR ^4^. This problem can largely be overcome by the bind/wash preconditioning step used in PCA (see methods section). Guanidinium thiocyanate is classified as a hazardous substance with corresponding transport restrictions, which may prevent its use in a non-laboratory setting. Therefore, as an alternative, and to reduce the necessary pipetting steps and simplify the workflow, we also tested inactivation and lysis of the viral particles in the sample by heat treatment (80 °C for 10 min). The efficacy of both treatments to inactivate SARS-CoV-2 was confirmed by plaque assay. There was no significant difference in the results of the PCA reactions with either treatment (data no shown).

Using a dilution series of SARS-CoV-2 viral particles grown on Vero E6 cells and inactivated with AVL buffer and ethanol we successfully detected 10e^5^ copies/µl of viral target RNA in 5 min 32 sec and 10e^1^ copies/µl in 17 min 39 sec (Fig 1A). Probit analysis showed the limit of detection (LOD) ^5^ to be at 4.9 copies/µl (95% confidence interval 3.8 – 8.2 copies/µl) (Fig. 2). A positive detection is defined as the measured fluorescence crossing the threshold fluorescence. The time it takes to reach this threshold (time to threshold - ttt value) is inversely proportional to the target copy number in the sample. Thus, the ttt value in PCA is analogous to the ct (threshold cycle) value given in qPCR. However, in contrast to qPCR, where a 10-fold dilution of target copy number is associated with an increase in ct value of 3.3, the ttt value in PCA does not follow such a strictly linear correlation with target copy number, particularly at low target concentrations (Fig. 1A). Therefore, PCA has to be regarded as a semiquantitative, rather than quantitative technology.

**Fig 1.**
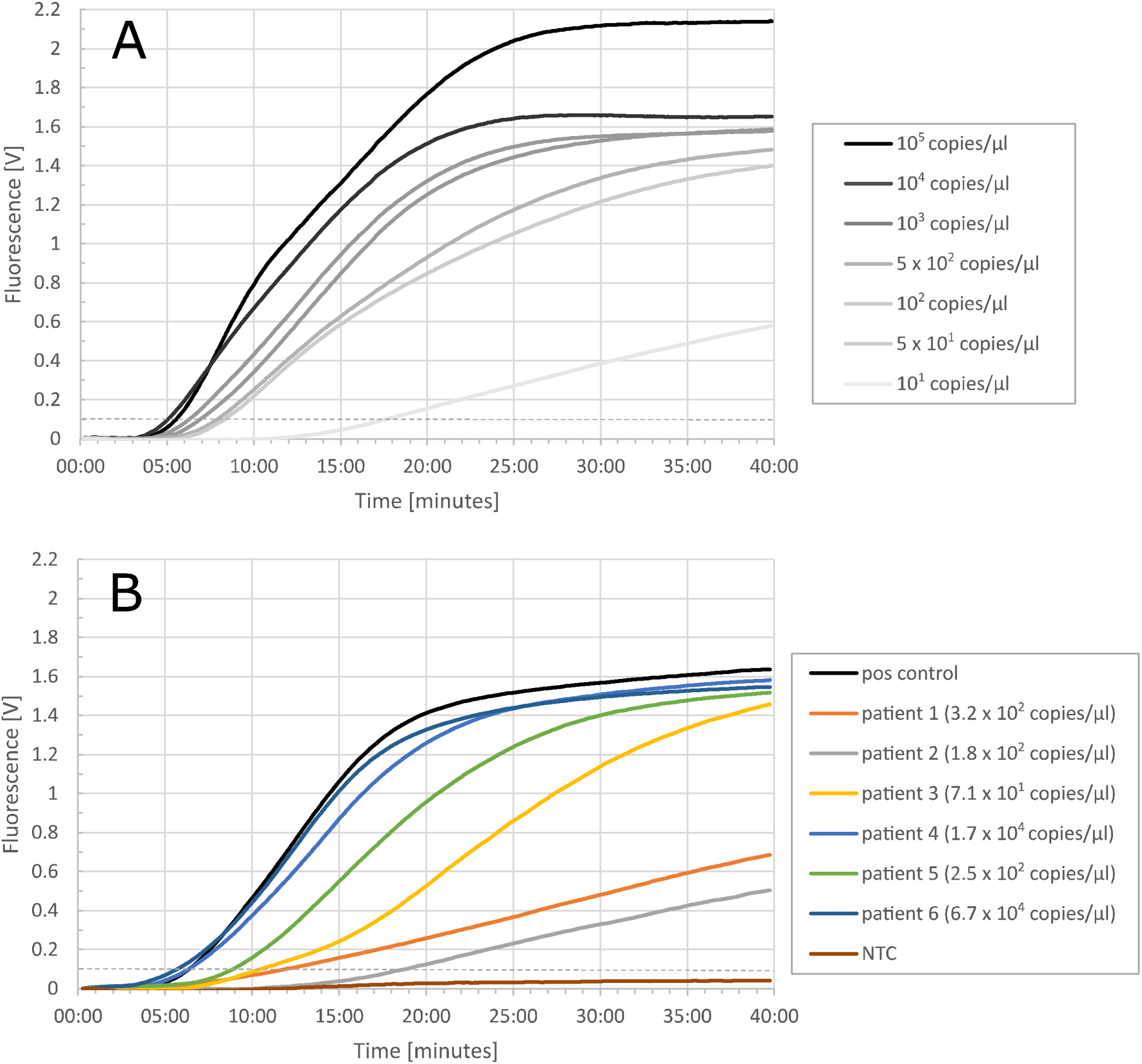
PCA performed with A) a dilution series of virus culture B) clinical samples. Dashed line denotes threshold fluorescence.

**Fig 2.**
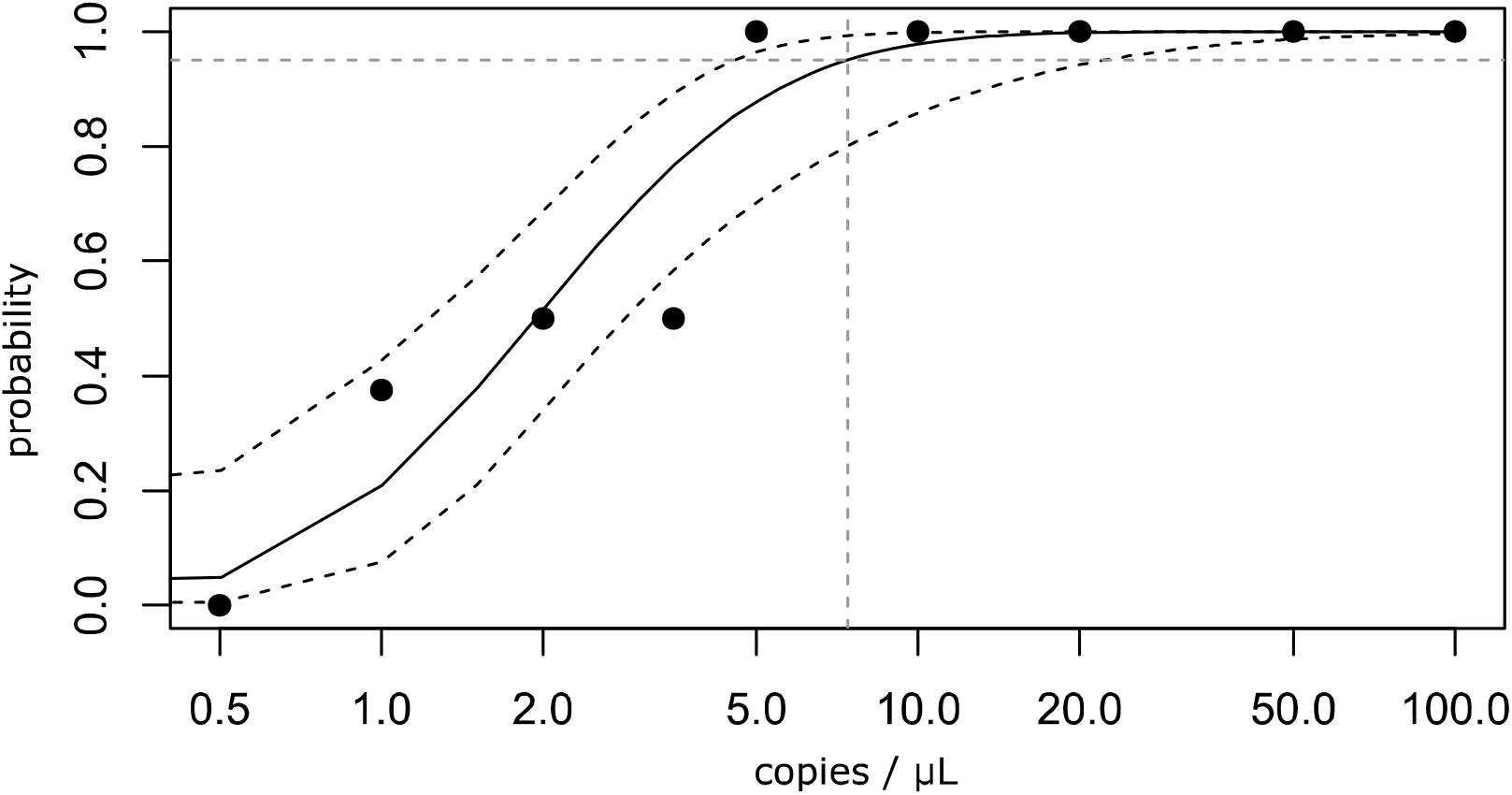
Probit analysis was done in replicates of eight using SARS-CoV-2 inactivated with 4 volumes of AVL buffer. Viral RNA was immobilized on the wires in two rounds of bind/wash. The limit of detection is 4.9 copies/µl (3.8 – 8.2 copies/µl, 95 % confidence interval). Extrapolated to the target copy number per PCA reaction, this equates to 443.7 copies (341.1 – 738.0 copies, 95% confidence interval). Dashed black lines denote the 95 % confidence interval.

We then tested 83 clinical samples that had been confirmed to be positive for SARS-CoV-2 using RT-qPCR targeting the E gene. A representative graph of the PCA results with these samples is shown in Fig. 1B. The concentration of target RNA in the 83 clinical samples ranged from 2.6 x 10^5^ to 7.4 x 10^0^ copies/µl. This corresponds to RT-qPCR ct values between 17.95 and 32.53. Out of the 83 samples 74 (89.2 %) tested positive in PCA, with ttt values ranging from 5 min 28 sec to 24 min 26 sec (Fig. 3). 100 % (n=32) of the samples with a RT-qPCR ct value <25 (corresponding to 1.6 x 10^3^ copies/µl) tested positive with PCA.

**Fig. 3.**
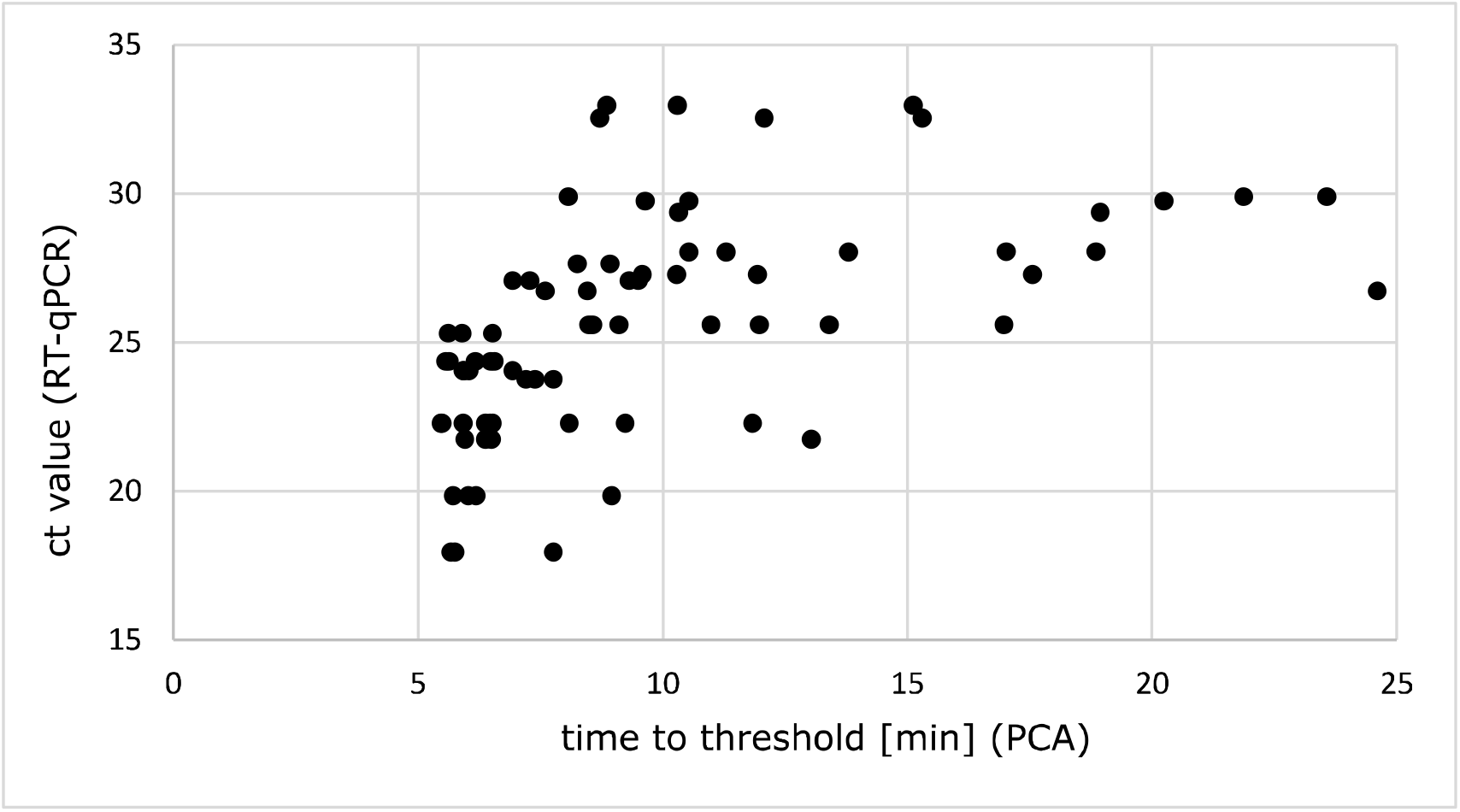
Comparison of the performance of PCA and standard RT-qPCR using clinical sample material (swab samples in universal transport medium).

To test diagnostic specificity, we used 40 clinical samples that had been confirmed to be negative for SARS-CoV-2 by RT-qPCR. All reactions were negative.

To test analytical specificity, we used a panel of 31 viral and bacterial pathogens that are relevant for differential diagnosis of respiratory infections (listed in table 2). All reactions were negative.

**Table 2.** Specificity panel to test for cross-reactions.

|  |  |
| --- | --- |
| 1 | *Acinetobacter baumannii* |
| 2 | *Bacillus anthracis* |
| 3 | *Burkholderia cepacia* |
| 4 | *Burkholderia mallei* |
| 5 | *Burkholderia pseudomallei* |
| 6 | *Burkholderia thailandensis* |
| 7 | *Candida albicans* |
| 8 | *Citrobacter freundii* |
| 9 | *Eikenella corrodens* |
| 10 | *Enterobacter aerogenes* |
| 11 | *Enterococcus faecalis* |
| 12 | *Escherichia coli* |
| 13 | *Francisella tularensis tularensis* |
| 14 | *Haemophilus influenzae* |
| 15 | *Klebsiella pneumoniae* |
| 16 | *Legionella pneumophila* |
| 17 | *Moraxella catarrhalis* |
| 18 | *Neisseria meningitidis* |
| 19 | *Propionibacterium acnes* |
| 20 | *Proteus mirabilis* |
| 21 | *Pseudomonas aeruginosa* |
| 22 | *Serratia marcescens* |
| 23 | *Staphylococcus aureus* |
| 24 | *Staphylococcus epidermidis* |
| 25 | *Stenotrophomonas maltophilia* |
| 26 | *Streptococcus pneumoniae* |
| 27 | *Streptococcus pyogenes* |
| 28 | Hantavirus |
| 29 | MERS Coronavirus |
| 30 | Influenza A virus |
| 31 | Influenza B virus |

## Discussion

Fast diagnostics of COVID-19 is crucial for management of the pandemic. Currently, RT-qPCR, which takes several hours to complete, is still the gold standard. In recent months, however, a number of faster alternatives based on different platforms and technologies have come onto the market that deliver results within an hour or less. On the one hand there are isothermal PCR methods like LAMP (loop-mediated amplification) ^6–9^ or RPA (recombinase polymerase amplification) ^10^, some of them in combination with a CRISPR-cas-based detection ^11–14^. On the other hand, there are fully integrated cartridge systems like Cepheid’s GeneXpert (Cepheid, Sunnyvale, USA) and Abbott’s IDnow (Abbott, North Chicago, USA). Further to these nucleic acid based technologies, the first antigen tests in lateral flow format are beginning to emerge, e.g. Covid-19 Ag Respi Strip (Coris Bioconcept, Gembloux, Belgium) or SOFIA2 SARS Antigen FIA (Quidel, San Diego, USA).

Apart from speed, spreading SARS-CoV-2 diagnostics over several different technologies can also be advantageous for a very practical reason. During the peak of COVID-19 infections in March-May 2020, where diagnostics was based almost exclusively on RT-qPCR, there were global supply shortages in PCR reagents, RNA extraction kits and platform-specific plasticware. For management of the ongoing pandemic and potential future surges of infections, having alternatives to RT-qPCR can alleviate the supply problems.

The published isothermal PCR assays for SARS-CoV-2 slightly lag behind the sensitivity of RT-qPCR, with LODs of 5-40 copies/µl ^6–14^ in contrast to 1-5 copies/reaction seen with some RT-qPCR assays ^1^. The combination of isothermal PCR with a CRISPR-cas-based detection system does not seem to lead to a better sensitivity. However, it may improve specificity, since it requires two target specific sequence recognitions, i.e. primer binding and CRISPR guide RNA binding. A detection directly from swab samples without prior RNA extraction has been described for some isothermal PCR assays ^6^ although this leads to some loss of sensitivity. The PCA assay described in this study is similar to isothermal PCR assays in terms of sensitivity, but is superior in terms of time to result (within 15-20 min as opposed to 30-60 min for isothermal PCR).

Cartridge-based systems have the obvious appeal of ease-of-use, since a sample can be added directly to the cartridge without any prior treatment and then inserted into the analyzer. Abbott’s IDnow delivers a positive result in 5 min, a negative one in 13 min and claims a LOD of 125 copies/ml sample (although this sensitivity has recently been questioned ^15,16^), while Cepheid’s Gene Xpert can detect 10 copies/ml in 45 min, which rivals the sensitivity of RT-qPCR on any open system. The disadvantage of cartridge systems is sample throughput. While the IDnow analyzer is certainly small enough for field or point-of-care use, it can only analyze one sample at a time. The Gene Xpert system is available with 1 up to 80 modules, but the multi-module devices are too bulky for field or POC use. In comparison, the Pharos Micro PCA device can analyze up to eight samples simultaneously in a small, lightweight device. A further disadvantage of cartridge-based tests is that they are closed systems, i.e. the target, primer and probe sequences they use are not publicly available. With a fast-evolving virus genome, mutations can appear over time, potentially with a geographically localized accumulation of certain lineages, which can affect test sensitivity. In a closed system the end user has no possibility to account for that.

Lateral flow assays (LFAs) are a classic example of POC tests. They deliver a result in 15-20 min and usually do not require any sample processing or instrumentation (although reader devices are available for some LFAs to provide semi-quantitative results) and require only minimal training to perform. As such, they are ideally suited for use in the field or at POC. However, LFAs also have a notoriously bad analytical sensitivity and usually fall several orders of magnitude short of the LOD of PCR ^17^. The COVID-19 Ag Respi-Strip (Coris Bioconcept, Gembloux, Belgium), a lateral flow assay based on detection with gold particles, has a diagnostic sensitivity of 60 % (76.7 – 85.7 % for samples with a RT-qPCR ct value <25). The Sofia2 SARS Antigen FIA (Quidel, San Diego, USA), a fluorescence immuno assay has a diagnostic sensitivity of 80 %, with a LOD of 8.5 x 10^2^ viral particles/ml. In contrast to the Coris Bioconcept test, the Quidel test requires an analyzer to detect the fluorescence signal.

The herein described PCA test is similar to LFAs in terms of time to result and fieldability, but provides higher sensitivity than LFAs. Its sensitivity is comparable to that of isothermal PCR techniques, but does not quite achieve RT-qPCR levels. It is, however, faster than isothermal PCR and considerably faster than RT-qPCR. With regard to sensitivity, the LOD of 4.9 copies/µl (or 4.9 x 10^3^copies/ml) determined for PCA is well below the mean viral load/ml measured in 3303 patients across all age groups of 5.18 – 6.39 (platform LC480, Roche) and 4.90 – 6.12 (platform Cobas, Roche) ^18^. Viral load in swab samples in the first 5 days after onset of symptoms was found to range between 10^3^ – 10^9^ /swab ^19^. Based on these figures, the SARS-CoV-2 PCA assay is likely to be able to detect the vast majority of COVID-19 infections in clinical samples.

While RT-qPCR and associated RNA extraction require a lab environment, PCA can be operated outside stationary laboratory facilities, even in the field ^2^. We envisage PCA as a point-of-care test for healthcare professionals, potentially in combination with more complex and time-consuming confirmatory tests. SARS-CoV-2 PCA tests can be used for a fast analysis of individual samples at or near point-of-care facilities to allow identification and immediate action on smaller clusters of infection or, with multiple PCA devices running in parallel, at drive-through testing stations with an on-site capacity of up to 2000 tests per day per 10 devices. With this approach most contagious patients, which show highest viral loads before the onset of symptoms ^20,21^ and in the early phase of the disease ^19^, as well as potential “silent carriers” could be rapidly identified and quarantined.

## Data Availability

Data is available from the authors upon request

## Author contributions

KZ planned the study, analyzed the data and wrote the manuscript, MW carried out the experimental work and analyzed data, CK carried out experimental work, RE cultured SARS-CoV-2 and tested inactivation treatments, RE, KM, RW and KS supported data analysis and revised the manuscript

